# Screening power of STIR muscle MRI in Critical illness myo-neuropathy and GBS in the ICU

**DOI:** 10.1101/2021.07.07.21259996

**Authors:** Boby Varkey Maramattom

**Author notes:** **Ethics committee approval**; Aster Medcity Ethics committee 2020; approved.

## Abstract

**Introduction:** Critical illness myo-neuropathy [CIMN] or ICU acquired weakness [CIMN] is a common cause of weakness in ICU patients. It is diagnosed by clinical features, nerve conduction studies and muscle/nerve biopsies.

**Methods:** The MRI muscle STIR images of 7 patients with suspected CIMN and 7 GBS patients were reviewed.

**Results:** All 7 patients with CIMN showed diffuse muscle edema, predominating in the lower limbs. Only one patient with GBS showed abnormal MRI changes (14%) and MRI was normal in 86%. The sensitivity of MRI to detect CIMN was 100%, whereas the specificity was 85.7%. Thus, the positive predictive value of MRI in this situation was 87.5% and the negative predictive value was 100%.

**Conclusions:** Muscle STIR imaging may help to differentiate between CIMN and GBS and can modify the diagnostic algorithm of CIMN.

## Introduction

Weakness in critically ill patients is one of the leading causes of ventilator dependence in the ICU. An important group is Critical illness myopathy [CIM], critical illness polyneuropathy [CIP] or a mixture of the two; Critical illness myo-neuropathy [CIMN]. It is also termed ICU acquired weakness [ICU-AW]. CIMN is suspected when ventilator weaning becomes difficult. Neurological examination discloses flaccid quadriparesis and areflexia with relative sparing of the cranial musculature. Sensory impairment is difficult to determine clinically as patients can be encephalopathic secondary to multiple underlying issues.^1^ CIMN can present as early as 5-7 days within admission to the ICU.^2^

Risk factors for CIMN include the severity and duration of critical illness, hyperglycemia, use of neuromuscular blockers or steroids, and circulatory factors in sepsis that reduce muscle membrane or nerve excitability.

Diagnosis of ICU-AW includes a combination of Medical Research Council [MRC] muscle strength grading, nerve conduction, EMG studies, direct muscle stimulation [DMS], muscle ultrasound and muscle/ nerve biopsies. An MRC sum score of <48 is suggestive of CIMN. Direct muscle stimulation (DMS) provides a “nerve/muscle ratio,” [nerve stimulation/ muscle-stimulation CMAP amplitudes]; with ratios of >0.5 indicating a neuropathic process [CIP], and a ratio <0.5 suggesting a myopathic disorder [CIM].^3^

The early diagnosis of CIMN in patients is hampered by factors such as alteration in sensorium, poor patient cooperation, ICU associated electrical artefacts, anasarca, necessity for specialized equipment, trained electro-physiologists and invasive tests. Although there is no specific treatment for CIMN at present, early diagnosis can potentially alter the subsequent ICU course and differentiate between subgroups of CIMN who respond to specific therapeutic interventions in future clinical trials. We have previously published an algorithm for the evaluation of a weak patient in the ICU.^4,5^ Diagnostic criteria for CIMN are also available.^6^

Guillain-Barre syndrome [GBS] is a close differential diagnosis of CIMN. It may be difficult at times to differentiate between GBS and CIMN. Some patients with CIMN undergo neuroimaging to assess the CNS or spinal cord when they have an encephalopathy or possible spinal cord dysfunction. However, MRI has not been systematically used to assess muscle involvement in CIMN or GBS. Only case reports of MRI abnormalities in GBS have been published.^7^ We have tried to assess the screening power of muscle STIR MRI imaging between confirmed cases of CIMN and GBS.

## Materials and methods

This was a retrospective study conducted over a 4-year period from February 2017 to February 2021. All adult patients with ICU-AW and GBS seen at Aster Medcity over this 4 year were included in the cohort and all patients had been examined by the lead author. Out of 20 patients with confirmed CIMN, 8 patients underwent MRI whole body muscle MRI. Out of 77 patients with confirmed GBS, 7 patients underwent whole body muscle MRI imaging and these images were also analyzed. Whole body muscle STIR imaging [short-tau inversion–recovery sequences (TR/TE, 3000–3665 ms/15–35 ms; inversion time, 150 ms) were obtained along with routine cranio-spinal imaging. Axial STIR images of the pelvis and thighs were obtained when possible. The images were interpreted by a radiologist with specialization in musculoskeletal imaging. The clinical records of all 15 patients and histopathological slides were collected. Institutional ethics committee approval was obtained (Aster Medcity Ethics committee 2020; approved). All patients underwent a standardized evaluation of muscle strength assessment, nerve conduction studies (NCS), electromyography (EMG) and lumbar puncture after 7 days. 5 of 7 patients with CIMN underwent a muscle biopsy. Standard criteria for ICU-AW and the Brighton criteria for GBS were used for diagnosis.^8^

## Results

7 cases of CIMN, Male: female ratio (4:3) with a median age range of 59 years (range 36-74 years) and 7 cases of GBS with a median age range of 57 years (age range 47-72 years) and a male: female ratio (4:3) underwent whole body STIR MRI at a median range of 16 days (11-28 days) for CIMN and 17 days (12-24 days) for GBS. Subcutaneous edema was found in 5 out of 7 patients (71%). All patients with CIMN showed STIR muscle hyperintensities (100%) by the 3^rd^ week of illness, whereas only 1 patient with GBS (14%) showed muscle hyperintensities. The MRI changes in the GBS patient was restricted to the proximal lower limbs and occurred in an AMAN variant.

Albumino-cytological dissociation was noted in all 7 GBS patients and none of the CIMN patients. Muscle biopsy in all 5 patients with CIM showed findings consistent with critical illness myopathy. The sensitivity of MRI to detect CIMN was 100%, whereas the specificity was 85.7%. Thus, the positive predictive value of MRI in this situation was 87.5% and the negative predictive value was 100%.

## Representative case

An elderly year lady with hypertension, type 2 diabetes mellitus and coronary artery disease was admitted with altered sensorium and intubated. Trop I was elevated and low molecular weight heparin was started. On day 7, she was conscious, quadriparetic [UL 1/5 power, LL 0/5], areflexic and had severe muscle tenderness in both thigh muscles. Her CRP was 33 mg/dl, CPK was 266 and CSF showed 3 cells with elevated protein [75mg%]. MRI brain revealed sulcal subarachnoid hemorrhage [SAH]. Her upper limb power improved to grade 4, however she had persistent thigh pain and paraparesis. NCS showed a sensori-motor axonopathy. MRI thigh on day 12 showed extensive muscle changes. [Panel C,D,E] Muscle biopsy showed diffuse atrophy of muscle fibres with angulated and shrunken fibres with peripheral nuclei with scattered foci of endomysial and perivascular chronic inflammatory infiltrate and vacuolization, consistent with CIM [non-necrotising cachectic type].

## Discussion

CIMN patients demonstrated extensive muscle edema on MRI which involved the lower limbs and pelvic muscles more than the upper limbs. Deep muscles (Obturator muscles & Ilio-psoas) which are difficult to assess by conventional techniques such as EMG or muscle biopsy also showed edema on MRI. Subcutaneous edema was found in CIMN patients (71%) reflected fluid imbalance secondary to the underlying critical illness. Our groups were comparable in terms of age and time to MRI imaging. With the use of targeted MRI sequences, we could demonstrate striking muscle abnormalities in all cases of CIMN by the 3^rd^ week of illness. The majority of GBS patients (86%) did not have any changes on muscle MRI. Only the patient with a severe AMAN variant showed proximal lower limb changes due to severe denervation

60% of patients with sepsis develop early neuromuscular and cardiac electrophysiological abnormalities on NCS and EKG [low SNAPs and CMAPs, reduced QRS complex amplitudes and increased QRS duration] within 72 hours of ICU admission. Of this cohort, 50% progresses to CIMN while the remainder show rapid improvement in electrophysiological parameters. This reversibility reflects temporary impairment of skeletal/ cardiac muscle membranes and nerve excitability and is attributed to circulatory factors in sepsis [such as Nitric oxide] which induce a nodopathy or paranodopathy.^9,10,11^ These inflammatory mediators inhibit mitochondrial activity and disrupt Na^+^/K^+^ ion channel pump function, resulting in Na^+^ influx, axonal depolarization and conduction failure.^9^ Later secondary Ca^2+^-mediated axonal degeneration supervenes. Thus NCS maybe abnormal only after 1-3 weeks or require repeated testing to confirm CIMN.^2,12,13^ Improvement on the other hand (clinical and electrophysiological) occurs over several months.^14^

When critically ill patients may undergo MRI for evaluation of an encephalopathy, muscle imaging is a logical extension of this diagnostic modality. MRI muscle changes in neuromuscular disease primarily occur due to muscle edema, mass lesions, fatty replacement or atrophy. The first two occur in subacute processes, whereas fibrosis and atrophy suggest chronic muscle disease. Muscle edema is seen in inflammatory myopathies, infectious myositis, radiation therapy, sub-acute denervation and other muscle diseases.^15,16,17^ Of these, inflammatory myopathies, rhabdomyolysis or vasculitis result in extensive and generalised muscle edema on MRI. Muscle edema is thought to be caused by intramuscular fluid shift, vascular congestion or perfusion alterations related to denervation or muscle inflammation. Following nerve injury or neuropathies, muscles innervated by that nerve undergo denervation and uniform edema. Edema is reflected as STIR hyperintensity in the affected muscles as early as 2-4 days after denervation although it can take upto 2-4 weeks for MRI abnormalities to manifest.^18,19 When^ effective re-innervation occurs, the MRI changes normalize. If re-innervation does not occur, it progresses to muscle atrophy with fatty infiltration in about 6 months.^20^ Unlike restricted muscle denervation following focal nerve injury, CINM is a unique entity characterized by diffuse denervation and muscle injury with corresponding scattered changes on MRI.

Neuromuscular imaging now incorporates multiple MRI sequences including T1W, T2W, STIR sequences, chemical shift imaging (CSI), diffusion weighted & diffusion tensor imaging (DWI/DTI), perfusion imaging, contrast imaging and MR neurography to differentiate muscle pathology.^17^ Performing all these sequences adds a considerable amount of time in a critically ill patient. Using only STIR sequences shortens MR imaging times and can be performed with concurrent cranio-spinal imaging.

Limitations of our study include a small sample size and the timing of MRI. Our patients could undergo an MRI only by the 3^rd^ week. The timing of MRI muscle imaging is crucial to pick up acute changes and we could not obtain MRI early in the course of the illness to see if it could contribute to therapeutic decisions.

The diagnosis of CIMN is often delayed until specialist neurologic opinion is obtained. MRI muscle imaging complements electrophysiological testing and also helps in targeting muscles for biopsy. Therefore, the addition of a STIR muscle MR imaging protocol in critically ill patients can expedite the diagnosis of CIMN. Furthermore, MRI STIR imaging permits a more objective diagnostic tool that is easier to interpret, for all physicians involved in the care of critically ill patients. The incorporation of MRI muscle imaging in the diagnostic evaluation of weakness in the ICU may be useful. [Chart 1]

**Chart 1.**
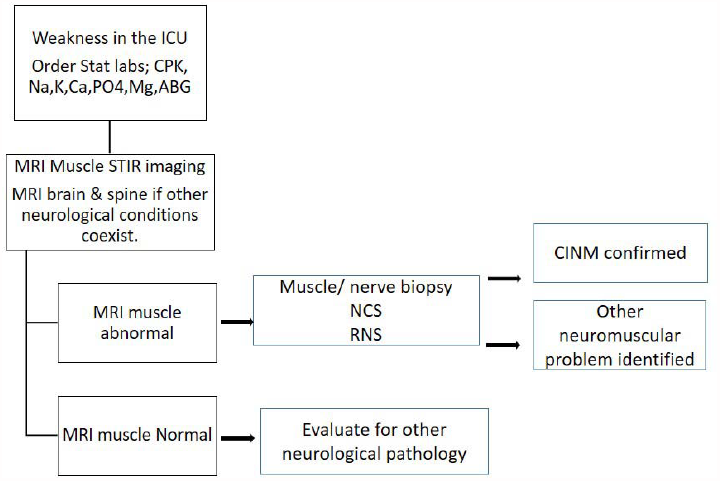
Algorithm for the evaluation of a weak patient in the ICU.

Future directions include the use of newer MRI sequences to differentiate CIMN subtypes as well as the use of MR neurography and DTI to assess concurrent CIP. Whether the use of muscle MRI earlier in the course of the illness helps in speeding up therapeutic decision is yet to be explored.

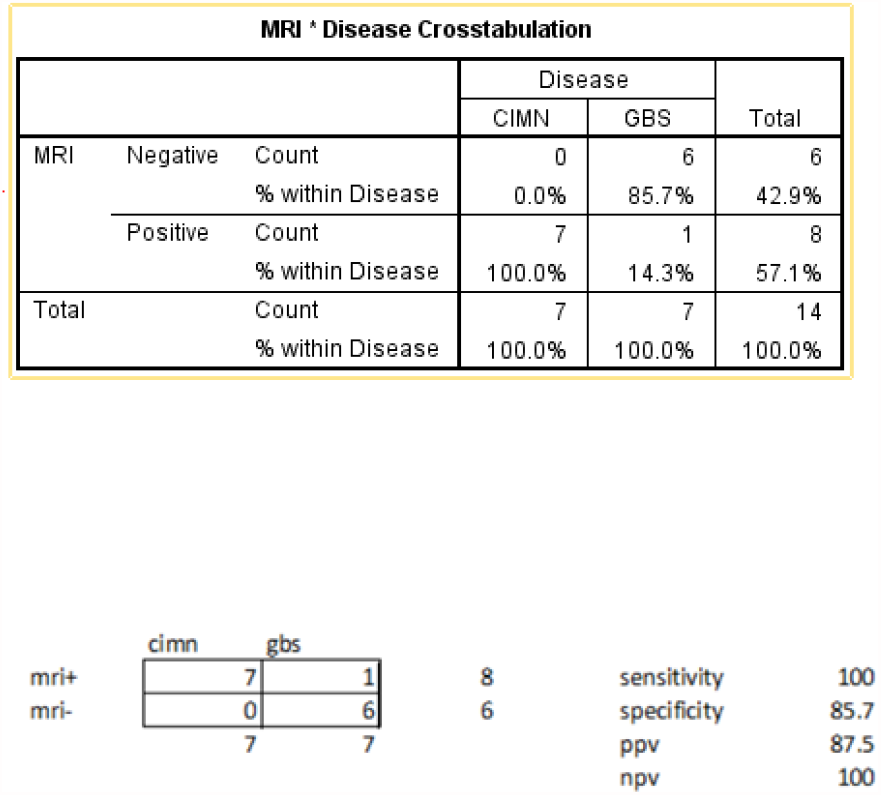

**Table 1.**
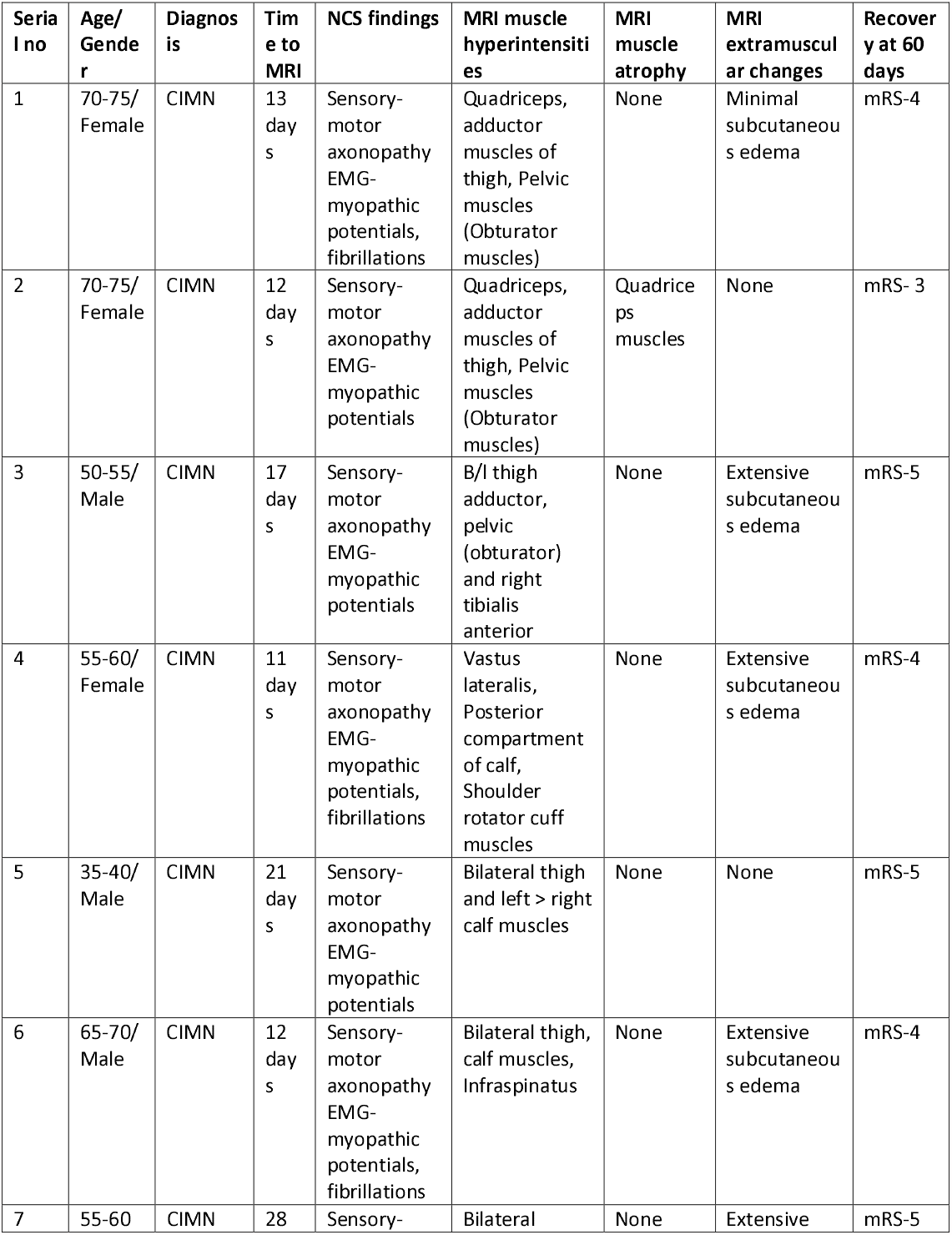
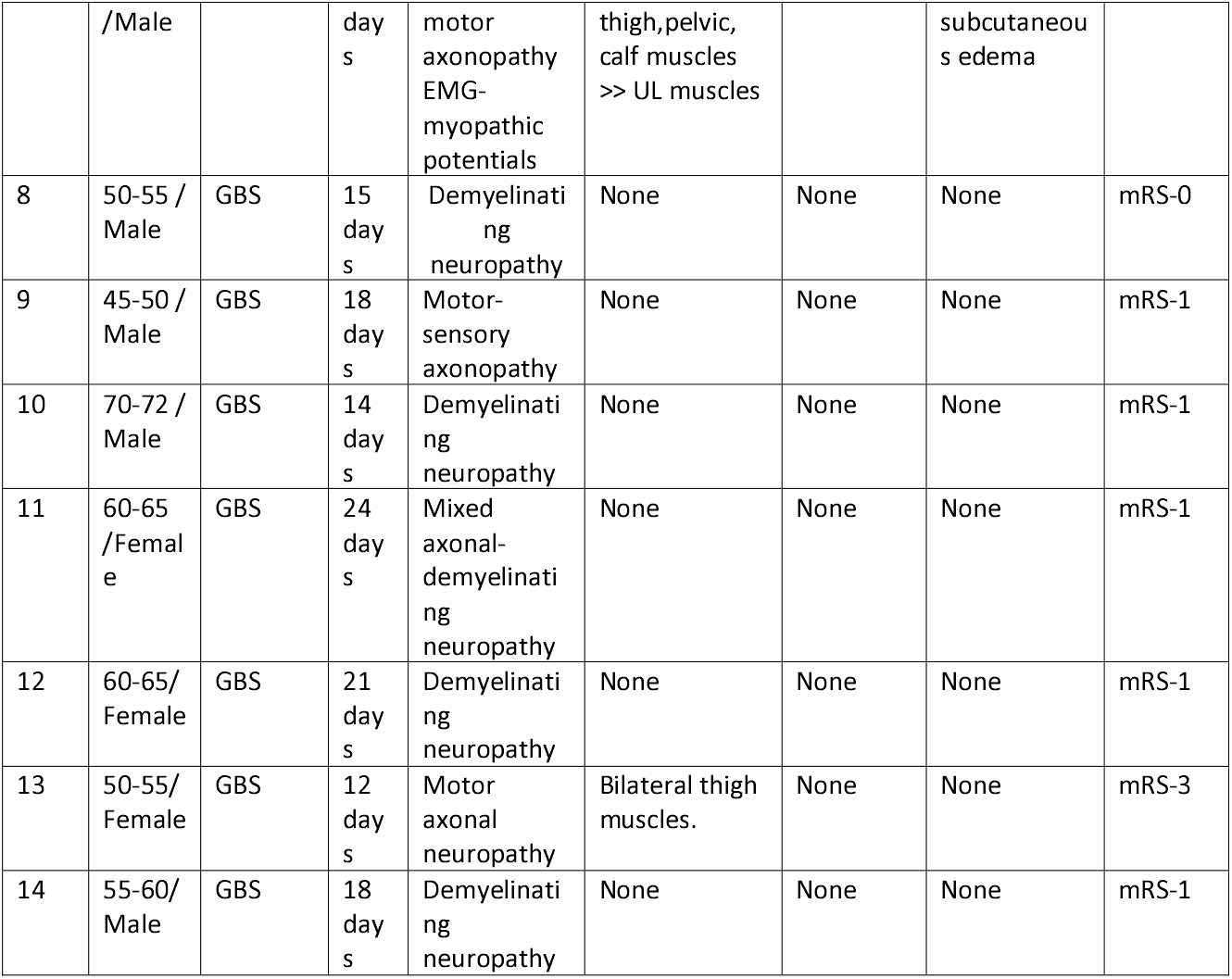
MRI abnormalities in patients with CIMN and GBS.

## Data Availability

Data are available on request

---

**Figure 1.**
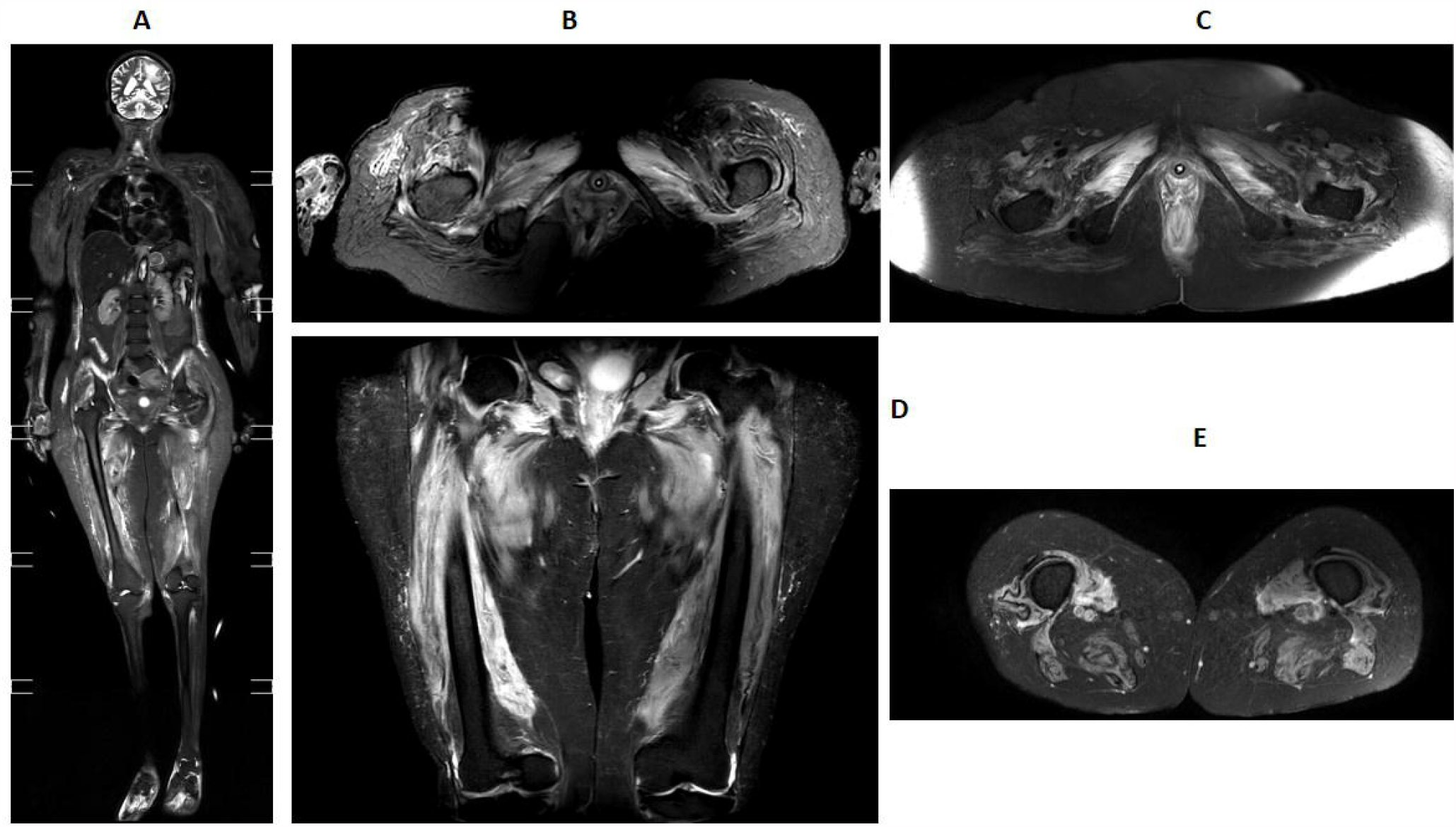
Panel A ;MRI Coronal STIR, Panel B;Axial STIR images of Patient 1. Panel D;Coronal STIR images of thigh. Panel C & E; Axial STIR images of thighs; Patient 2

**Figure 2.**
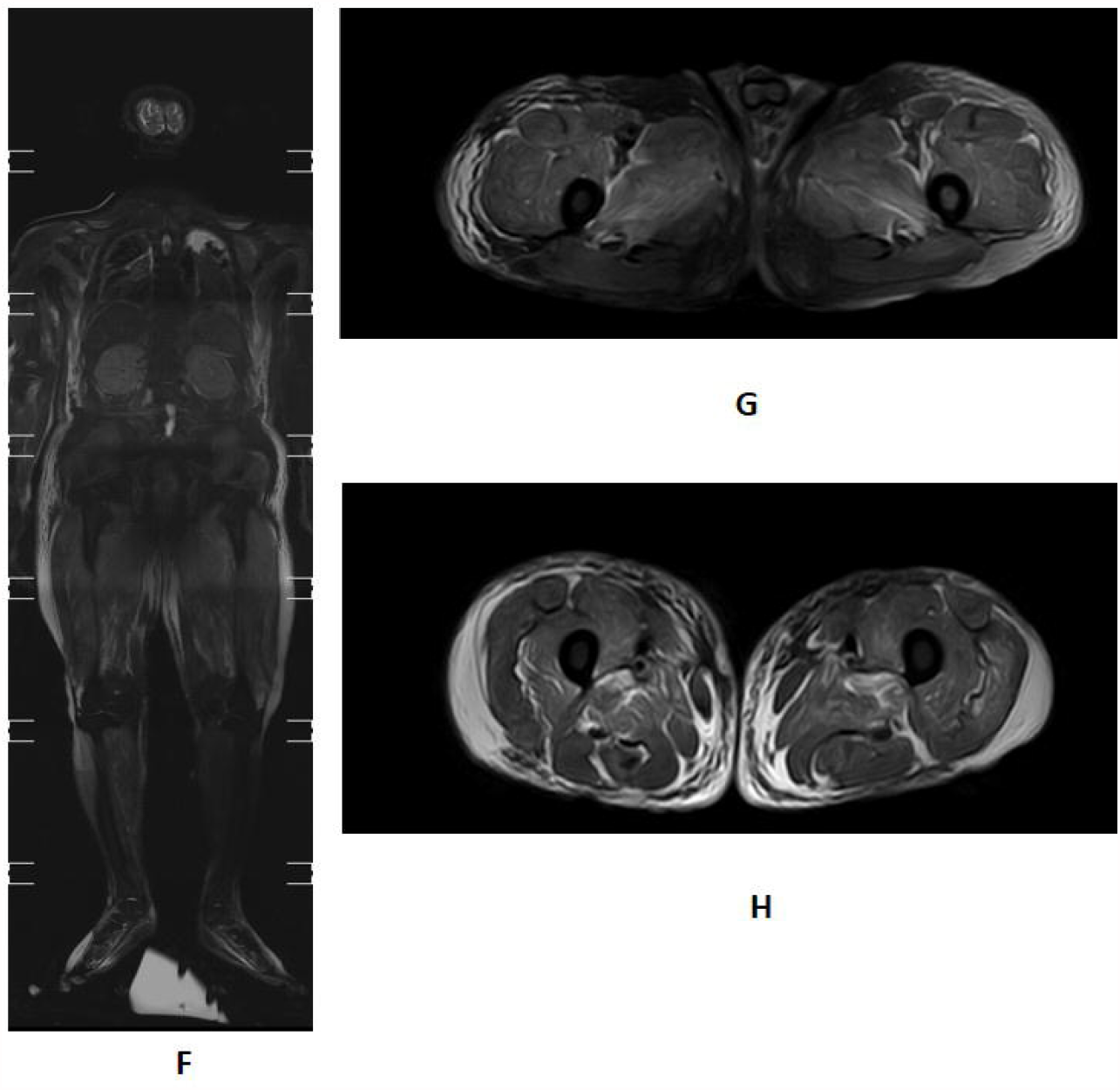
Panel F,G,H; Coronal and axial STIR images of patient 3.

**Figure 3.**
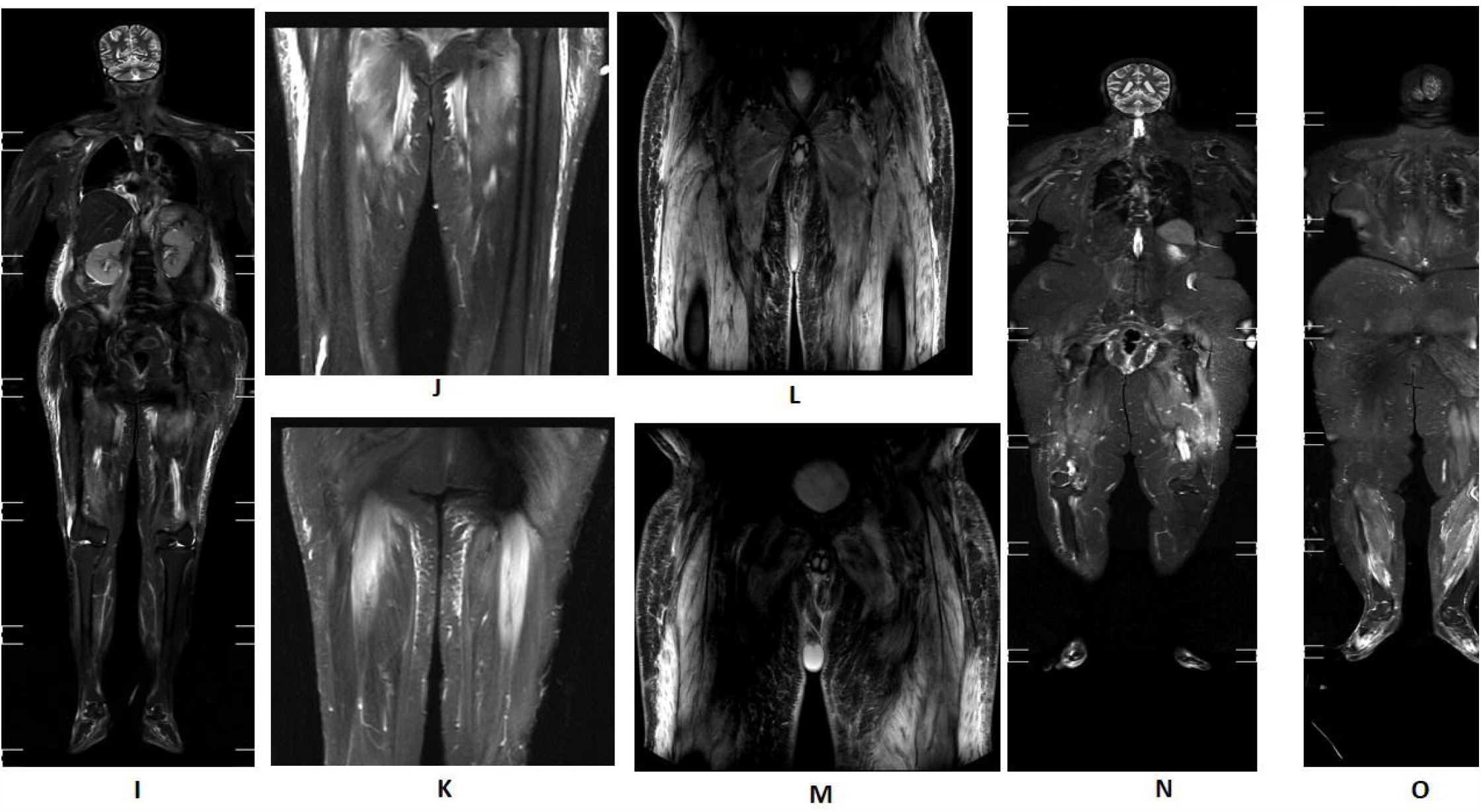
Panels I,J,K; Patient 4, STIR Coronal whole body [I], Coronal anterior thigh [J], Posterior thigh [K] Panels L,M; Patient 5; Coronal images through anterior and mid thigh Panels N,O; Patient 6; Coronal STIR images through anterior [N] and posterior planes [O]

**Figure 4.**
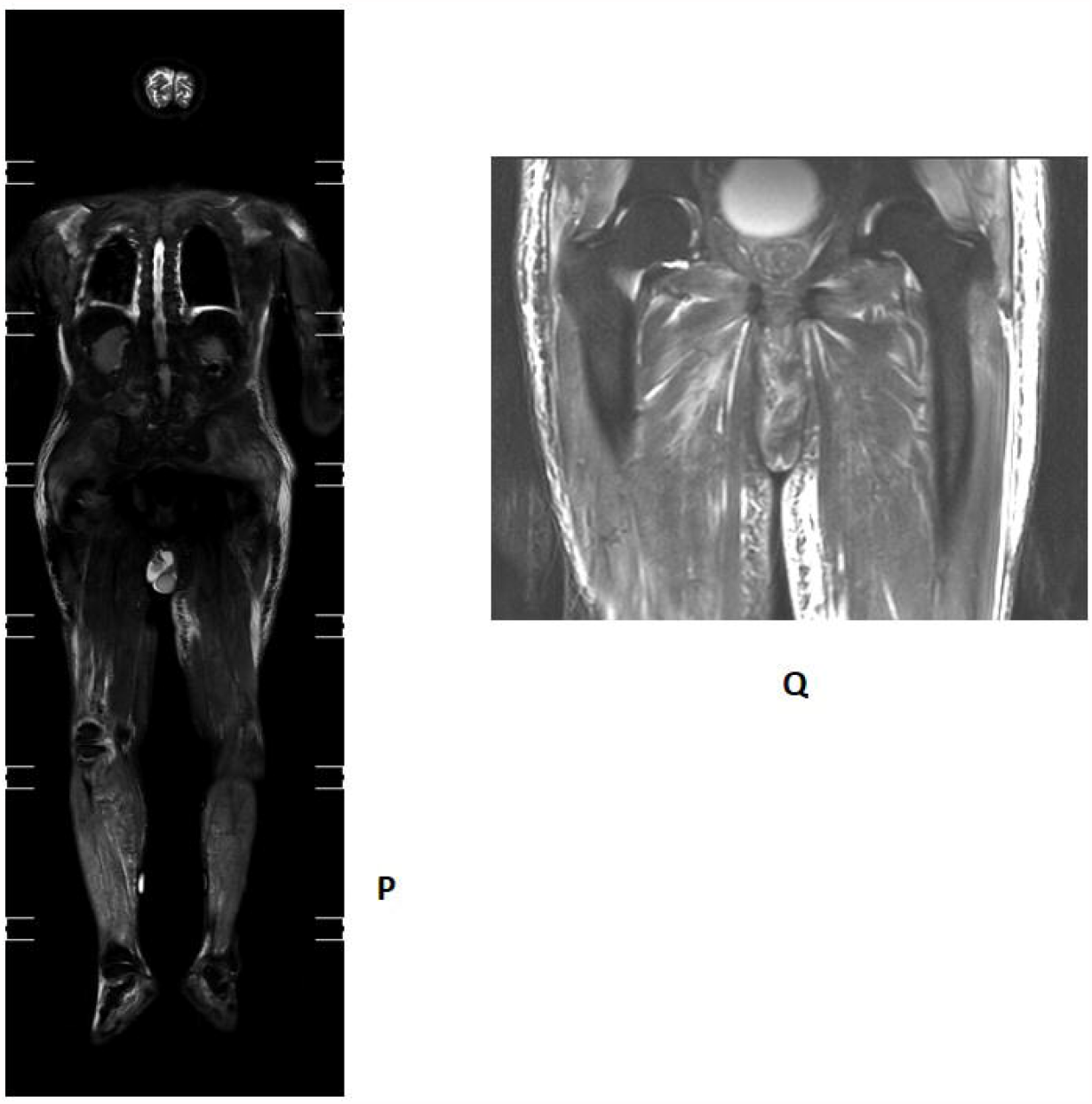
Panel P; Coronal STIR MRI image, Panel Q; Coronal STIR image through thigh.

## Notes

### Competing Interest Statement

The authors have declared no competing interest.

### Author Declarations

Ethics committee approval; Aster Medcity Ethics committee 2020; approved

